# Modified Alliance-Focused Training with Doubling as an integrative approach to improve therapists’ competencies in dealing with alliance ruptures and prevent negative outcome in psychotherapy for depression. Study protocol of a randomized controlled multicenter trial

**DOI:** 10.1101/2025.01.12.25320410

**Authors:** Antje Gumz, Denise Kästner, Laurence Reuter, Carmen Martinez Moura, Klarissa Ehlers, Anne Daubmann, Catherine F. Eubanks, J. Christopher Muran, Timothy Anderson, Ramona Stöckl, Georg Schwanitz, Lena Stegemann, Lana Rohr, Ulrike Willutzki, Frank Jacobi, Antonia Zapf

**Affiliations:** Psychologische Hochschule Berlin, Germany, Department of Psychosomatics and Psychotherapy, Berlin, 10179; 00491635026408; Psychologische Hochschule Berlin, Germany, Department of Psychosomatics and Psychotherapy, Berlin; Institute of Medical Biometry and Epidemiology, University Medical Center Hamburg-Eppendorf, Germany; Derner School of Psychology, Adelphi University, Garden City, New York, USA; Derner School of Psychology, Adelphi University, Garden City, New York, USA; Psychotherapy Research Program, Icahn School of Medicine at Mount Sinai; Department of Psychology, Ohio University, Athens, Ohio; Clinical Trial Office, Charité Universitätsmedizin Berlin, Germany; University Witten/Herdecke, Department of Psychology and Psychotherapy, Witten, Germany; Psychologische Hochschule Berlin, Germany, Department of Clinical psychology and Psychotherapy, Berlin, 10179; 004930209166220

**Keywords:** Alliance-Focused Training, depression, psychotherapy, randomized controlled trial, therapeutic alliance, alliance ruptures

## Abstract

**Introduction:** Alliance ruptures constitute a high risk of premature treatment termination and poor psychotherapy outcome. The Alliance-Focused Training (AFT) is a promising transtheoretical approach to enhance therapists’ skills in dealing with alliance ruptures.

**Methods and analysis:** To evaluate the effectiveness of Modified AFT with doubling (MAFT-D), a randomized, patient and evaluator-blinded, multicenter trial was designed comparing MAFT-D (delivered to trainee therapists and supervisors) and psychotherapy training/ treatment as usual (TAU) for therapists (n=120) and their patients with depressive disorders (n=240). A total of 16 cooperating centers, each offering either cognitive-behavioral or psychodynamic psychotherapy training, will contribute to recruitment. Stratifcation by center (both for therapists and patients) and hence therapeutic approach (cognitive-behavioral vs. psychodynamic psychotherapies), and by comorbid personality disorder (yes vs. no, for patients) will be carried out. The two hierarchically ordered primary hypotheses are: In MAFT-D compared to TAU, a stronger reduction of depressive symptoms and a lower rate of patient dropout is expected from baseline to 20 weeks after baseline. Follow-up assessments are planned at 35 weeks, 20 months, and 36 months post-baseline to evaluate the persistence of effects. Secondary patient-and therapist-related outcomes as well as predictors, moderators, and mediators of change will be investigated. Mixed models with repeated measures will be used for the primary analyses.

**Ethics and dissemination:** Ethical approvals were obtained by the institutional Ethics review board of the main study center as well as by review boards in each federal state where one or more cooperating centers are located (secondary votes). Following the Consolidated Standards of Reporting Trials statement for non-pharmacological trials, results will be reported in peer-reviewed scientific journals and disseminated to patient organizations and media.

**Trial registration number:** DRKS00014842 (German Clinical Trial Register).

**Strengths and limitations of this study:**

- This large randomized controlled multicenter trial aims to deepen our understanding of alliance-focused trainings and holds great potential to substantially advance evidence-based psychotherapy training across therapeutic approaches.
- The study is designed to assess the effects of MAFT-D (delivered to trainee therapists and supervisors) on both therapists and, most importantly, on their patients.
- Involving multiple psychotherapy training institutes not only ensures generalizability, but may also facilitate transfer into clinical training and practice.
- By focusing on depression while stratifying for comorbid personality disorders, the study also stands to clarify potential differential effects of MAFT-D depending on patients’ psychopathological characteristics.
- The supervision frequency (one session per four therapy sessions), aligned with local real-world practices, and the primary measurement point (20 weeks), chosen for comparability with other trials, together pose a risk of insufficient dose, with effects potentially becoming apparent only at later follow-ups.

## Background

Depressive disorders are among the most common mental disorders (1) and the leading cause for disability worldwide (2). The course of the disorder is often recurring or chronic and the consequences for the individual and the society can be severe (1). The guideline-recommended treatments for depression are pharmacotherapy, psychotherapy or a combination of both (depending on severity and chronicity). Cognitive-behavioral and psychodynamic psychotherapies (CBT and PDT) are evidence-based and comparably effective (1, 3–5). However, unsatisfactory outcomes in depression treatments are frequent (up to 41% of patients do not reliably improve, up to 1/3 drop out (6–8).Thus, factors closely linked to outcome and dropout need to be addressed. Here, the evidence clearly points to the importance of common factors (relevant across therapeutic approaches), especially the therapeutic alliance (9–12).

Alliance ruptures (i.e., periods of tension or breakdown of the collaborative relationship between patient and therapist) inevitably occur in the course of any psychotherapy (13). Across various therapeutic approaches, there is an evergrowing wealth of evidence establishing a link between alliance ruptures and treatment outcome, whereas the therapists’ ability to repair ruptures has been demonstrated to improve outcome and to prevent dropout (14–27). However, therapists often fail to notice ruptures or lack the skills to deal with them constructively (13, 28–32).

Consequently, it is of crucial importance to improve the competence of therapists in dealing with alliance ruptures. This is in line with current recommendations according to which psychotherapy trainings should give more emphasis to common therapy principles like improving the alliance (33–35). Despite the common occurrence of alliance ruptures and the serious implications resulting from failures to resolve them, the curricula of psychotherapy training do not systematically address the topic as of yet.

Various therapist trainings focus on therapeutic relational skills and/or the process of alliance rupture and repair. Among them, the most prominent is Alliance-Focused Training (AFT, (36, 37)), which follows a transtheoretical approach and is grounded in an evidence-based model developed by Safran, Muran, and Eubanks for successfully resolving alliance ruptures (13). Its main objective is to strengthen three skills: 1) to sensitively notice alliance ruptures, 2) to tolerate difficult affects, and 3) to meta-communicate about them in a helpful way. Video-recorded sessions and role plays are fundamental elements of AFT (37–39). Building on our pilot study (funded by the Heigl Foundation), we made several modifications to the training, the most significant of which was the incorporation of the psychodrama technique of doubling after role plays to enhance the focus on affective communication and to foster a secure, non-judgemental atmosphere in both training and supervision (40, 41) (details see section “Experimental intervention”).

Previous research emphasizes the potential of therapist training programs targeting the therapeutic alliance in general, and of AFT specifically (20, 42, 43). A meta-analysis of empirically supported relationship factors (44) describes the systematic resolution of alliance ruptures as one (out of three) of the most promising relationship elements that warrant further research. Two recent studies demonstrated an effect of AFT on therapists’ skills (42, 43). Evidence on patient outcomes appears promising regarding the reduction of dropout rates, but remains unclear in terms of symptom improvement (20, 22). Overall, the evidence on the effectiveness of trainings focusing on the alliance seems still rather limited. The studies included in the existing reviews and meta-analyses (20–22) were highly heterogeneous with respect to the investigated patient populations, treatment lengths, scope and content of the training intervention. Moreover, the sample size of most studies was small. Promising results were found with respect to patients with depressive disorders (evidence base: study 1: n=31 intervention group (IG) 1; n=34 IG 2; n=38 control group (CG); 8 sessions problem solving therapy (45); study 2: n=11 IG, n=11 CG, 16 sessions cognitive therapy (46)).

Despite the central role of relationship problems among patients with depression (47–49) and the high proportion of negative or insufficient therapeutic outcome, there is no large-scale trial investigating the benefits of trainings with a focus on the therapeutic alliance in depression. More studies are needed to investigate the effectiveness of AFT and to link the increase in therapist competence to outcome (50).

Against this background, our study intends to investigate the benefits of our modified form of AFT (MAFT-D) vs. psychotherapy training as usual (TAU) for patients with depressive disorders. The decision to investigate patients with depressive disorders is based on the facts that a) to date there is no large-scale study investigating the effects of alliance-focused trainings in depression, b) depressive disorders are highly prevalent and debilitating, c) interpersonal problems are key factors in models on the development and maintenance of depression (48, 49), d) negative outcome in depression treatments is particularly frequent (6–8). The project investigates CBT and PDT therapists, because of the transtheoretical nature of AFT, the great significance of common factors, and the urgent need for evidence-supported methods in psychotherapy training. Training and therapy under routine conditions represents the reference for efficacy and safety for the experimental intervention, because CBT and PDT are established evidence-based treatments for depression, and psychotherapy training at the state-approved institutes can also be considered as the current gold standard.

Additionally, the project intends to investigate mediators of MAFT-D effectiveness as well as further evidence-based mediators of of change, as well as predictors and moderators of therapy success.

## Methods

### Study design

Patients, their trainee therapists and therapists’ supervisors will take part in the randomized controlled multicenter trial. The trial intends to investigate the benefits of MAFT-D vs. TAU for patients with depressive disorders, stratified by the cooperating center (psychotherapy training institute), subsequently by therapeutic approach (CBT vs. PDT) and comorbid personality disorder (SCID-5-PD, yes vs. no), as well as their trainee therapists, stratified by cooperating center (psychotherapy training institute) and therapeutic approach (CBT vs. PDT). Therapists and supervisors belonging to the intervention group will receive MAFT-D. Therapists and supervisors belonging to the the control group will receive/conduct psychotherapy training as usual (TAU). Figure 1 illustrates the study design. The trial is registered at the German Clinical Trial Register (DRKS00014842).

**Figure 1.**
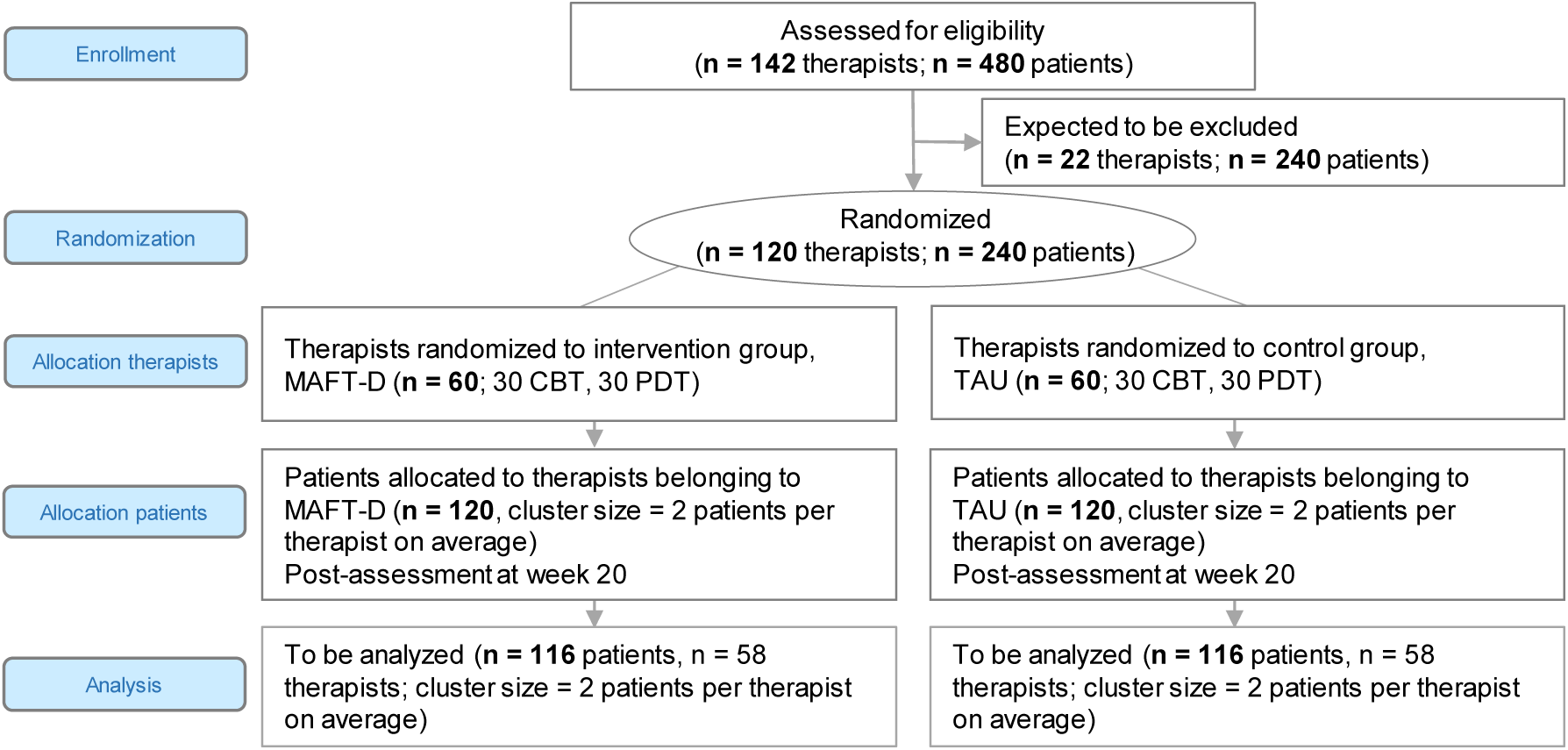
Trial design. CBT: Cognitive-behavioral therapy; PDT: Psychodynamic therapy; MAFT-D: Modified Alliance-Focused Training with doubling; TAU: Psychotherapy Training as usual.

### Hypotheses and research questions

The two hierarchically ordered primary hypotheses are: In MAFT-D compared to TAU, a stronger reduction of patient-rated depressive symptoms (first primary endpoint) and a lower rate of patient dropout (second primary endpoint) is expected from baseline to 20 weeks after baseline.

Secondary patient-related hypotheses are: In MAFT-D compared to TAU, stronger reduction of patient-rated depressive symptoms after 35 weeks, and 20 and 36 months, lower rate of dropout at week 35, and month 20 and 36, stronger reduction of observer-rated depressive symptoms, and patient-rated anxiety, somatic complaints, personality structure deficits, interpersonal problems, and quality of life after 20 and 35 weeks, and 20 and 36 months.

Secondary therapist-related hypotheses are: In MAFT-D compared to TAU, stronger increase in observer-rated interpersonal skills after 35 weeks, and therapist-rated therapeutic skills, trait-like relational manner in therapy, emotional suppression, alexithymic traits (difficulties identifying and describing feelings) and satisfaction with supervision after 20 and 35 weeks, and 20 and 36 months.

The hypotheses regarding mediators of MAFT-D effectiveness are: MAFT-D generates more favorable outcomes via an improved therapeutic alliance and increased therapist skills to deal with alliance ruptures. Corresponding mediator variables are: 1) therapeutic alliance (51), 2) ratio of unresolved/ resolved alliance ruptures (22), 3) convergence of patients’ and therapists’ alliance ratings over time (14, 52, 53), 4) interventions referring to the therapeutic relationship in the here and now (13), 5) nondirective supportive therapist’s techniques (54, 55), 6) therapists’ competence in session (empathy) (56), 7) adherence to MAFT-D (13, 22).

Subsidiary research questions concern predictors, moderatos, and mediators of a) therapeutic macro and micro outcomes (e.g., dropout, alliance), b) the effectiveness of MAFT-D (e.g., presence of a personality disorder or therapeutic orientation as moderators of MAFT-D effectiveness), c) observer-rated interpersonal therapeutic skills and skills development, and d) the successful resolution of alliance ruptures.

### Methods Against Bias

#### Randomization and stratification

Therapists are randomly assigned by a 1:1 ratio to receive MAFT-D vs. TAU stratified by study center and approach (CBT/PDT). Patients are randomly assigned to MAFT-D vs. TAU stratified by study center, subsequently by approach (CBT/PDT), and comorbid personality disorder (SCID-5-PD, yes vs. no). Block randomization with variable block length is used. Patients are assigned to therapists appropriate to the random group. Randomizations will be performed centrally by Prof. Zapf’s group, University Medical Center Hamburg Eppendorf. Allocation is done via the electronic case report forms (implemented in secuTrial®).

#### Blinding

Therapists will be aware of allocation, patients will be blinded. Interviewers, raters and persons conducting the statistical analyses will be blinded. There will be strictly separate role assignments in the study team (raters/interviewers vs. unblinded study team members). Raters/ interviewers will work physically separated from unblinded staff.

#### Allegiance bias

Principal investigator (PI), research group members and national or international collaborators are PDT, CBT or systemic therapists, recruiting centers are from both therapeutic approaches, statisticians have no affiliation to a therapeutic approach.

#### Contamination bias

All MAFT-D-supervisors and -therapists will be asked to treat the contents of the intervention highly confidential and to give a standardized response to respective questions. MAFT-D-therapists will receive a two-staged informed consent (second stage: description of MAFT-D, after randomization). Part of the intervention (workshop) will be offered to TAU-therapists at the end of follow-up. Supervisors in the intervention group will document the total number of cases discussed during each supervision session and and the number in which MAFT-D was used. The supervisors are also asked retrospectively about the perceived usefulness of individual MAFT-D elements. Additionally, the number of hours therapists spent engaging with crucial elements of the intervention - such as observing role plays, participating in role plays, watching video-recorded therapy sessions of other therapists, and reviewing their own video-recorded sessions - is included in the “basic characteristics, experience, and practice” section of the assessment battery. Therapists complete this section at all fixed measurement points (BA, Post, FUP 1, 2, 3). Differences between therapists in the intervention group and the control group regarding these items will be analyzed. To estimate the extent of contamination bias, an adherence measure score (Beth Israel Fidelity Scale (57)) will be applied to the video-recorded therapy sessions and analyzed as dependent variable using a linear mixed model.

#### Other biases

Data monitoring will be conducted by the Clinical Study Office Charité Berlin (CTO). In order to minimize potential bias due to video-recordings, all therapy sessions will be recorded. The number of theoretical hours and supervision sessions completed will be compared for MAFT-D- and TAU-therapists. The use of additional treatments of the patients is monitored and analyzed (Client Sociodemographic and Service Receipt Inventory (58)). Therapists’ training preferences are analyzed as efficiency modifiers. To assure generalizability, we use a multi-center design. To assess selection bias, we will document ineligeble persons among patients (basic demographic information, reason for non-participation). Cases of withdrawal who allow to use their data will be compared with participants who do not drop out regarding baseline differences. We will document reasons for study discontinuation. Incentives are used to reduce missing values in other outcomes (e.g., 150€ dropout post-assessment).

### Inclusion and exclusion criteria for patients

#### Inclusion criteria

a) Agreement to start CBT or PDT outpatient therapy, weekly 50 min sessions; b) Age ≥ 18 years; c) Present DSM-5 diagnosis of depressive and related disorders (major depressive disorder - single or recurrent episode; persistent depressive disorder, other specified depressive disorder). Operationalization: Structured Clinical Interview for DSM-5 Disorders, Clinician Version, SCID-5-CV (59, 60), conducted by trained raters. The SCID-5-CV shows excellent to satisfactory interrater-relaibility, via phone and face-to-face (61, 62); d) Reaching or exceeding the cut-off value indicating moderate depression severity. Operationalization: Beck Depression Inventory II, BDI-II (63, 64), score ≥ 20 (1, 65); e) Signed informed consent form

Rationale for the criteria: Depression is the most common mental disorder in Germany, associated with considerable costs (66). Psychotherapy is recommended for patients with moderate or severe depression (1). Interpersonal problems are key factors in models on the development and maintenance of depression (47–49, 67).

#### Exclusion criteria

a) Current or past diagnosis of a psychotic or bipolar disorder; b) Substance dependency disorder (current or during the last 12 months); SCID-5-CV at timepoint of screening; c) Acute suicidal plans at timepoint of screening or suicide attempts within the last 6 months; modified (last six months) Colombia Suicide Severity Rating Scale (CSSR-S) (68), which is a reliable and valid instrument for identification of suicide risk (68, 69); d) Psychopharmacotherapy other than antidepressants; change of antidepressant regimen during the previous month or during study participation; modified Client Sociodemographic and Service Receipt Inventory (CSSRI) (58); which is one of the most widely used instruments for a standardized assessment of health care utilization (including medications; medication classification will be based on ATC-codes) (58); e) Planned concurrent psychological treatments (Self-help groups can be continued.); CSSRI; f) Insufficient command of German language, below level B2.

Rationale for the criteria: Severe comorbidities are excluded for reasons of patients’ safety. Concomitant treatments are limited to minimize performance bias. The number of exclusion criteria is kept low to ensure representativeness for routine care.

### Inclusion criteria for therapists and supervisors

#### Therapists

a) graduated psychologist or licensed physician; b) in advanced CBT or PDT training (with a license to practice outpatient therapy under supervision, which is usually obtained after 2 years of training); c) capacity to start treatment with two new patients, d) capacity to visit one of the workshops offered for study therapists.

#### Supervisors

a) approved by state and cooperating institute, b) no scheduling conflicts (e.g., workshop dates, regular dates for group supervision), c) agreement to decline requests for supervision from study therapists of the other condition during the study period.

There are no exclusion criteria for therapists and supervisors.

### Cooperating study centers

The main study center is at Psychologische Hochschule Berlin where study coordination and data collection take place. The cooperating centers across Germany are state-approved, well established psychotherapy training institutes offering courses in individual outpatient psychotherapy (CBT or PDT). They support the recruitment of participants and are responsible for organization and carrying out of the study-independent regular therapy training and patient treatments. The following centers agreed to cooperate (alphabetically ordered, person responsible in brackets):

1. Akademie für Psychotherapie Erfurt (Prof. M. Geyer)
2. Berliner Akademie für Psychotherapie (Prof. F. Jacobi)
3. Centrum für Integrative Psychologie Bamberg (Dr. J. Siegl)
4. DGVT-Ausbildungszentrum Berlin (Dr. M. Rotter)
5. Dresdner Institut für Psychodynamische Psychotherapie (Dr. C. Schilling, Dr. S. Seifert)
6. ppt-Institut für Psychologische Psychotherapie und Beratung Berlin (Dr. L. Hauten, Dr. R. Spielberg, E. Stahl, S. Ulm)
7. IFT – Psychotherapeutische Ambulanz, München (S. Gmeinwieser)
8. Institut für Psychotherapie und Angewandte Psychoanalyse, Jena (Dr. U. Wutzler)
9. Leipziger Ausbildungsinstitut für Psychologische Psychotherapie und Universität Leipzig (Prof. C. Exner, Dr. S. Koranyi)
10. Regionalinstitut Sachsen der DGVT (K. Sturmeit)
11. Sächsisches Institut für Psychoanalyse und Psychotherapie (Prof. K. v. Klitzing, Dr. O. Krauß, A. Kazzer)
12. Universität Greifswald (Prof. E.-L. Brakemeier, M. Tewes)
13. Universität Witten/Herdecke und DGVT-Ausbildungszentrum Dortmund (Prof. U. Willutzki)
14. Weiterbildungsstudiengang Psychodynamische Psychotherapie, Klinik und Poliklinik für Psychosomatische Medizin und Psychotherapie, Universitätsmedizin der JGU Mainz (Prof. M. Beutel)
15. Saarländisches Institut für Tiefenpsychologisch fundierte Psychotherapie, Saarbrücken (Dr. E. Hahn, W. Bauer-Neustädter)
16. Zentrum für Ausbildung in Psychologischer Psychotherapie, Friedrich-Alexander-Universität Erlangen-Nürnberg (Prof. M. Berking, Dr. K. Zierhut)

Other centers will be involved in the event of recruitment difficulties.

### Recruitment

Recruitment will take place at the cooperating study centers (see above). Additional centers will be involved in the event of recruitment difficulties. Recruitment started in July 2024 and is ongoing.

The standard group size for group supervision is four participants which applies to the study and also to routine therapy training. Consequently, units of eight therapists must usually be recruited per institute. Some institutes allow smaller group sizes. However, a group consisting of only two participants on a long-term basis (more than 4 months) is considered a protocol violation. Combining study therapists from different training institutes at one location in a supervision group is permitted. The groups can also be filled with non-study participants. Changing the format from group to individual supervision is generally not permitted within the context of the study. Recruitment methods for therapists include project promotion through e-mail, advertisment in lectures/ seminars. Patients are recruited from the regular outpatient pool. Recruitment period is 21 months for patients and 16 months for therapists. Patients, therapists and supervisors will receive incentives for study related expenses (assessments, video recordings, travel).

### Patient treatments

Basic conditions of treatments will be equal between MAFT-D and TAU. Patients will receive weekly 50-minute sessions (regular frequency and length). Post-assessment will take place at week 20, first follow-up at week 35, 2nd and 3rd follow-up after 20 and 36 months. Treatments can be short- or long-term depending on patient’s and therapist’s agreement and the sessions granted by payers. All sessions will be video recorded. Therapists will treat two study patients on average. The costs of treatments are fully covered by statutory health insurances in Germany.

### Psychotherapy training

TAU- and MAFT-D-therapists will continue to participate in their regular training courses. Group supervisions of treatments will be carried out by experienced supervisors (usual frequency, one supervision session per four therapy sessions, in person, additional individual supervision is permitted in accordance with the regulations of the specific institute in the event of crises or for questions regarding the report for the health insurance company) either with MAFT-D specific focus or according to the usual procedure. The supervision groups are fixed and in order to participate the therapist must have already started at least one study therapy. Group supervisions will occur monthly or bi-weekly with patients being discussed for varying amounts of time (10 to 50 minutes per case). Supervisors will receive regular supervision fees. The hours of the workshop for the MAFT-D-group will replace other non-mandatory courses of the regular training.

### Experimental intervention

Therapists randomized to MAFT-D will receive a two-day workshop (approx. 15 teaching units, 11.5 hours) followed by group supervision carried out by supervisors who will also be trained in MAFT-D. The initial workshops will take place in person (for therapists and supervisors separately). Central elements are a theoretical introduction (150-180 min.) and role plays based on individual case examples (approx. 45 min. per participants). The number of lecturers depends on the overall group size (one lecturer and max. eight therapists per role play subgroup). In the case of unexpected absences (e.g., illness) and for therapists who are unable to attend at the originally planned workshop dates, catch-up workshops will be offered. Catch-up workshops consist of smaller groups and are shorter (approx. 10 teaching units, 7.5 hours), but are more intense due to more frequent participation in role plays.

AFT is a transtheoretical training concept initially developed by Muran, Safran, and Eubanks (36) focusing on alliance ruptures and aiming at enhancing therapists’ interpersonal skills to a) sensitively recognize alliance ruptures, b) tolerate negative affects, and c) meta-communicate using video-recorded sessions, role plays, and mindfulness exercises (13, 36, 40, 41). We have translated the AFT into German and tested and modified it in pilot studies (37, 40, 41). Our modifications concern (31, 70, 71): a) strong standardisation of the supervision process, b) verbalization of the affective experience exclusively via “doubling” (psychodrama technique) immediately after the role plays, c) structured focussing on therapist‘s own contribution (vulnerabilities, biographical relationship experiences; based on pre work (72, 73)), d) active role of supervisors in practicing meta-communication (“prompting”). The modification were made, because in our experience, there is a tendency in supervision groups to rationalize, intellectualize and cognitively exchange concepts and strategies, which can also conceal difficulties in opening up affectively (resistance to the perception of subjective or objective countertransference (72, 74)) or “hiding” of alexithymic tendencies (i.e. difficulties in perceiving and expressing emotions; (75)). The modifications aim to enhance the focus on the perception and verbalization of emotions, minimize intellectualizing after role plays, and foster a more non-judgmental atmosphere within the supervision group. Supervision follows the following standardized procedure (duration approx. 45 minutes per patient): A) Mindfulness exercise (to increase awareness and openness to one’s own emotional experience), 3-4 minutes; B) Introduction of the patient and characteristics of the therapeutic relationship with this patient (B1: verbally, B2: video sequence from last therapy session, B3: “screenplay” of an alliance rupture), 8-10 minutes; C) First role-play and “doubling” (C1: Supervisee in therapist role, supervision group member in patient role, C2: “Doubling”, psychodrama technique, to deepen the affective insight and enable access to blind spots), 7-9 minutes; D) Second role-play and “doubling” (D1: Supervisee in patient role, further supervision group member in therapist role, D2: “Doubling”, D3: Supervisor asks group members who were in the patient role how they felt), 10-13 minutes; E) Third role-play and own contributions (E1: Supervisee in therapist role, tries out new behavior, practises meta-communication, uses suggestions from the supervisor – prompting, E2: Supervisor asks what the rupture may have to do with supervisee’s vulnerabilities, whether own biographical issues may have been ‘triggered’, which feelings may be difficult to bear due to personal experiences), 6-8 minutes.

Supervisors belonging to MAFT-D will receive a two-day workshop before start of study therapies (same contents as for therapists but separately), followed by a monthly two-hour online training to refine MAFT-D-specific supervision skills and discuss questions.

To ensure consistency across centers, MAFT-D-therapists of different centers will be trained together. The same applies to supervisors. There will be a fixed group of lecturers (experienced psychotherapists and experienced in conducting MAFT-D-specific workshops and supervision).

### Sample size calculation, number of study participants

### Depressive symptoms

There are no prior studies that closely align with our study in terms of study population, intervention, and measurement instruments. In our view, the chosen reference (Bambling et al., 2006; (45)) represents the best available compromise, as it examines a population of depressed patients, employs an alliance-focused supervision approach, and uses the BDI-II as a measurement instrument. Accordingly, we assume a higher reduction in BDI-II after 20 weeks in MAFT-D compared to TAU of 4.0 BDI-II units with a standard deviation of 7.3 BDI-II units. To detect such an effect with a type I error of .05 (two-sided hypothesis), a power of .90, an intra-cluster correlation coefficient of .05, and a cluster size of 2 patients per therapist on average, we need 76 therapists and 152 patients in total (38 therapists and 76 patients per group, PASS 15, tests for two means in a cluster-randomized design).

#### Treatment dropout

We expect that MAFT-D will reduce the treatment dropout rate after 20 weeks from 30% to 12.5%, corresponding to an odds ratio of 3.0. The expected dropout rate of 30% for TAU is a conservative estimate based on a recent meta-analysis (35% dropout) (7) and numbers in our own clinic (n= 3.357 depressed patients; 33% dropout). Previous studies on different alliance-focused training approaches reported a difference in treatment dropout rates of 17.5% or greater (Bambling et al.: IG1=6.1%, IG2=3%, CG=35%; Constantino et al.: IG=0%, CG=27.3%; Muran et al.: IG=20%, CG1=37%, CG2=46%) (45, 46, 76). With identical design effects, 58 therapists and 116 patients per group (116 therapists, 232 patients in total) are needed to detect this difference (PASS 15, test for two proportions in a cluster-randomized design). We expect that 3% of the randomized sample are not part of the full analysis set (oriented on numbers from the trial of (77)) due to the lack of any data post-randomization (78). Thus, based on the larger sample size required for dropout, a sample of 120 therapists and 240 patients in total has to be enrolled.

Based on previous psychotherapy trials (77, 79), we expect a loss to follow-up of 15% (of therapists with their patients) at post-assessment in BDI-II measurement. With the planned number of sample size and the assumed loss to follow-up, we can prove the assumed effect. We assume a non-compliance rate for the (per protocol) BDI-II analysis (e.g., failure to complete 12 sessions within 20 weeks, change in antidepressant medication, use of additional treatments) of 35%.

We expect that 20% of patients and therapists decline study participations and 15% of therapist and 50% of patients do not meet inclusion criteria. Consequently, 178 therapists and 600 patients need to be initially invited in order to assess eligibility in 142 therapists and 480 patients, in order for 120 therapists and 240 patients to participate in the study and finally analyze 116 therapists and 232 patients.

### Endpoints, variables, instruments

#### Primary endpoints

Change in patient-rated depressive symptoms will be measured using the total score of Beck Depression Inventory (BDI-II, change=difference from baseline to 20 weeks after baseline) (65, 80) which is one of the most commonly used self-rating scales to assess depressive symptoms and is particularly recommended for monitoring the severity of depressive symptoms over time in clinical populations (1). Items are rated from 0 ‘not present’ to 3 ‘severe’. The total score is the sum of all 21 items (0–63). Good psychometric properties (reliability, validity, sensitivity to change) were demonstrated for the German version (65).

Premature therapy dropout is a dichotomous variable determined by patient interview. It is defined as the patient’s decision to end therapy contrary to the initial agreement with the therapist, excluding premature terminations unrelated to therapy quality, such as those due to serious illness of the patient or therapist, or moving to a different city. In case patients cannot be reached, interviews will be conducted with the therapist, and the patient is classified as dropout.

#### Patient-related secondary endpoints

- Change in patient-rated depressive symptoms (BDI-II total score) and therapy dropout at the remaining measuring times.

Relating to difference from baseline to 20 and 35 weeks, 20 and 36 months after baseline:

- Observer-rated depressive symptoms (Hamilton Rating Scale for Depression, GRID-HAM-D (81, 82), total score of the 21-item version). Excellent interrater-reliability (ICC, intra-class correlation = .95), Cronbach’s α: .78.
- Patient-rated anxiety (Generalized Anxiety Disorder Scale-7, GAD-7 (83), total score, rated on a 4-point Likert scale, 0 ‘not at all’ to 3 ‘nearly every day’). Cronbach’s α: .92, good test-retest reliability (ICC = .83) (83).
- Patient-rated somatic symptoms (Patient Health Questionnaire-15, PHQ-15 (84), total score, ratings of problems in the last four weeks, 0 = ‘not bothered at all’ to 2 ‘very bothered’). Cronbach’s α: .80, validity was proven (85).
- Patient-rated personality structure deficits (Operationalized Psychodynamic Diagnosis-Structure Questionnaire, short version, OPD-SQS (86), total score, 12-items assessing personality functioning, rated on a 4-point Likert scale, 0 ‘does not apply at all’ to 4 ‘does apply completely’). Cronbach’s α: .89 (86).
- Patient-rated interpersonal problems (Inventory of Interpersonal Problems-Revised, IIP-32 (87), total score, 32 items scored on a 5-point Likert scale). Adequate internal consistency was demonstrated. Cronbach’s α of the subscales: .68 to .90 (88).
- Patient-rated quality of life (World Health Organization Quality of Life, short version, WHOQOL–BREF (89), 5-point Likert scale, 1 ‘not at all’ to 5 ‘completly/to an extreme amount’. We will use two dimensions: physical and psychological quality of life. Chronbach’s α of the subscales: .57 to .88.

#### Therapist-related secondary endpoints

- Difference in observer-rated interpersonal skills from baseline to week 35 after baseline of the first patient (Facilitative Interpersonal Skills Performance Test, FIS (90–94), total score, eight items measuring general interpersonal skills based on verbal responses to six video clips displaying challenging therapy situations - verbal fluency; hope; persuasiveness; emotional expression; warmth, acceptance and understanding; empathy; alliance bond capacity and rupture-repair responsiveness), 1 ‘low level’ to 5 ‘high level’ of skills). The German FIS version shows good interrater-reliability (total score ICC = .81). Cronbach’α: .95 to .96 (91, 93).
- Therapist-rated retrospective satisfaction with supervision at week 20, 35, months 20, 36 after baseline of the first patient (modified version of the Client Satisfaction Questionnaire-8 (95), total score, measuring satisfaction with supervision instead of treatment, CSQ-8-mod (40). Cronbach’α: .87 to .93, good concurrent validity for the total score (96).

Relating to difference from baseline to 20 and 35 weeks, 20 and 36 months after baseline (all therapist rated):

- Trait-like relational manner and therapeutic competence score (relational manner - trait section (35 items) and skills change index section (10 items) of the Adapted Trainee Current Practice Report which is originally used in the SPRISTAD trial and currently undergoing psychometric evaluation (97)).
- Emotional suppression (Emotional Regulation Questionnaire subscale score, ERQ (98), 4 items, 1 ‘totally disagree’ to 7 ‘totally agree’). Cronbach’s α suppression subscale: .74.
- Alexithymia (Toronto Alexithymia Scale, TAS-26 (99), toal score and the difficulties a) identifiying und b) describing feelings subscales, 5-point likert scale, 1 ‘does not apply at all or not true at all’ to 5 ‘completely true’). Cronbach’s α: .80 to .86 (99).

#### Variables to assess potential mediators for M-AFT-D effectiveness

- Average level of therapeutic alliance across sessions (German short revised version of the Working Alliance Inventory, WAI-SR (100–102), assessed after each session, 12 items). Cronbach’s α for the total score: >.90. Convergent validity is established (101).
- Convergence of patient’s and therapist’s alliance ratings over time (patient and therapist versions of the WAI-SR, total scores of session ratings; see above).
- Ratio of unresolved/ resolved alliance ruptures (Post-Session-Questionnaire – modified version, PSQ-mod (103, 104). The first item asks whether a “problem or tension” occurred in the relationship between patient and therapist during the session. If answered with “yes”, the intensity of the rupture and the degree to which it could be solved is specified. Despite the simple nature of the items, there are clear indications of their validity (103, 104).
- Therapists’ usage of interventions referring to the therapeutic relationship in the here and now as well as usage of nondirective supportive techniques (mean of the categories “repeating, paraphrasing, summarizing”, „expression of emotional sympathy”, „validation”); Psychodynamic Intervention List, PIL, observer-ratings based on transcribed therapy sessions on the level of verbal statements (105), 0 ‘category characteristics do not apply at all’ to 5 ‘category characteristics completely apply’. Satisfactory reliability and convergent validity; interrater-reliabilities ICC “expression of emotional sympathy”: .69, ICC “validation”: .68, ICC “repeating, paraphrasing, summarizing”: .80, ICC “referring to the therapeutic relationship: .78, ICC topic “therapist”: .87 (55, 105).
- Therapists’ empathy (Empathy Scale (106)), total score, 10-items, observer-ratings of video-taped sessions, 0 ‘no agreement at all’ to 3 ‘very strong agreement’. Cronbach’s α: .86 (107).
- Therapists’ basic communication skills (Clinical Communication Skills Scale – short version, CCSS-S; (108), total score, 14 items, observer-ratings of video-taped sessions, 4-point scale ‘not at all appropriately’ to ‘entirely appropriately’. Reliability and validity was demonstrated (108).
- Therapists’ adherence to AFT, CBT, PDT (Beth Israel Fidelity Scale, BIFS, (57), observer-ratings of video-taped sessions, 12-items each (three subscales). Good psychometric properties, criterion validity and adequate inter rater agreement have been demonstrated repeatedly (109).

#### Variables for subsidiary research questions

- Mediators of change (patient-rated, Mediators of change in Psychotherapy Inventory, MoCPI (110)), after each session, 22 items and five subscales: quality of the therapeutic relationship (7 items), patients’ perception and expression of feelings and thoughts (5 items), patients’ becoming conscious of biographical experiences (4 items), gaining a new view on problems (4 items), patients’ self-disclosure (2 items). A five-factor structure with good psychometric properties could be found (110).
- Parenting behavior of therapist’s parents (German translation of the Measure of Parental Style Questionnaire, Fragebogen dysfunktionaler elterlicher Beziehungsstile, FDEB (111, 112), parenting style of father and mother, 15 items each, 1 ‘does not apply at all’ to 4 ‘does completely apply’). Cronbach’s α: .48 to .93 (112).
- Therapists’ personality traits (Big Five Inventory, short version, BFI-10 (113), measuring extraversion, agreeableness, conscientiousness, emotional stability, openness, 10 items). Cronbach’s α: .49 to .62 (114).
- Therapists’ interpersonal problems (IIP-32, see patient outcomes).
- Therapists’ sensitivity to aversive interpersonal behaviour (Interpersonal Sensitivities Circumplex Questionnaire, ISC-G (115), 40 items assessing the extent to which individuals are bothered by others’ interpersonal behaviors, 1 ‘not at all, never’ to 8 ‘extremely, always’). Cronbach’s α: .74 to .89.
- Therapists’ abilities to mentalize (Certainity About Mental States Questionnaire, CAMSQ (116), 20 items, 0 ‘never’ to 1 ‘always’, measuring the perceived capacity to understand mental states of the self and others. McDonald’s Ω: .88 to .91, good validity and test-retest reliability (116).
- Therapist’s dispositional fear of negative evaluation (Fear of Negative Evaluation Scale, FNE (117), German version - Skala Angst vor negativer Bewertung, SANB (118), 5 items, 1 ‘almost never applies’ to 4 ‘almost always true’). Cronbach’s α: .84 to .94.
- Therapists: Relational manner – state (97), 35 items, see therapist-related endpoints.
- Therapists’ perception of own competence and skills (Facilitative interpersonal skills, FIS, self rating; see therapist-related endpoints.
- Patients’ therapy expectations and therapy evaluation, PATHEV (119), 11 items, 1 ‘not true at all’ to 5 ‘completely true’, three subscales: improvement, fear of change, suitability. Chronbach’s α: .73 to .89.
- Patients’ early depressive symptom changes (pre to 5th session, BDI-II (65, 80), see above).
- Distinctive internal reactions (Impact message Inventory, IMI-C (120–122), observer-ratings, 64 items, 1 ‘not true at all’ to 4 ‘absolutely true’, dimensions: affiliation and dominance. Chronbach’s α: .84 to .89, interrater-reliability, ICC: .30 to .58.
- Planned for following funding period: Rupture Resolution Rating System, 3RS (123), observer-ratings, presence of a rupture and its clarity and intensity, 5-point Likert-scale. Interrater-reliability withdrawal rupture ICC: .85, confrontation ruptures ICC: .98 (124).
- Selected sessions will be analysed in terms of nonverbal behavior and linguistic variables.

### Measurement time points

The data collection is summarized in **Table 1**.

**Tab. 1:**
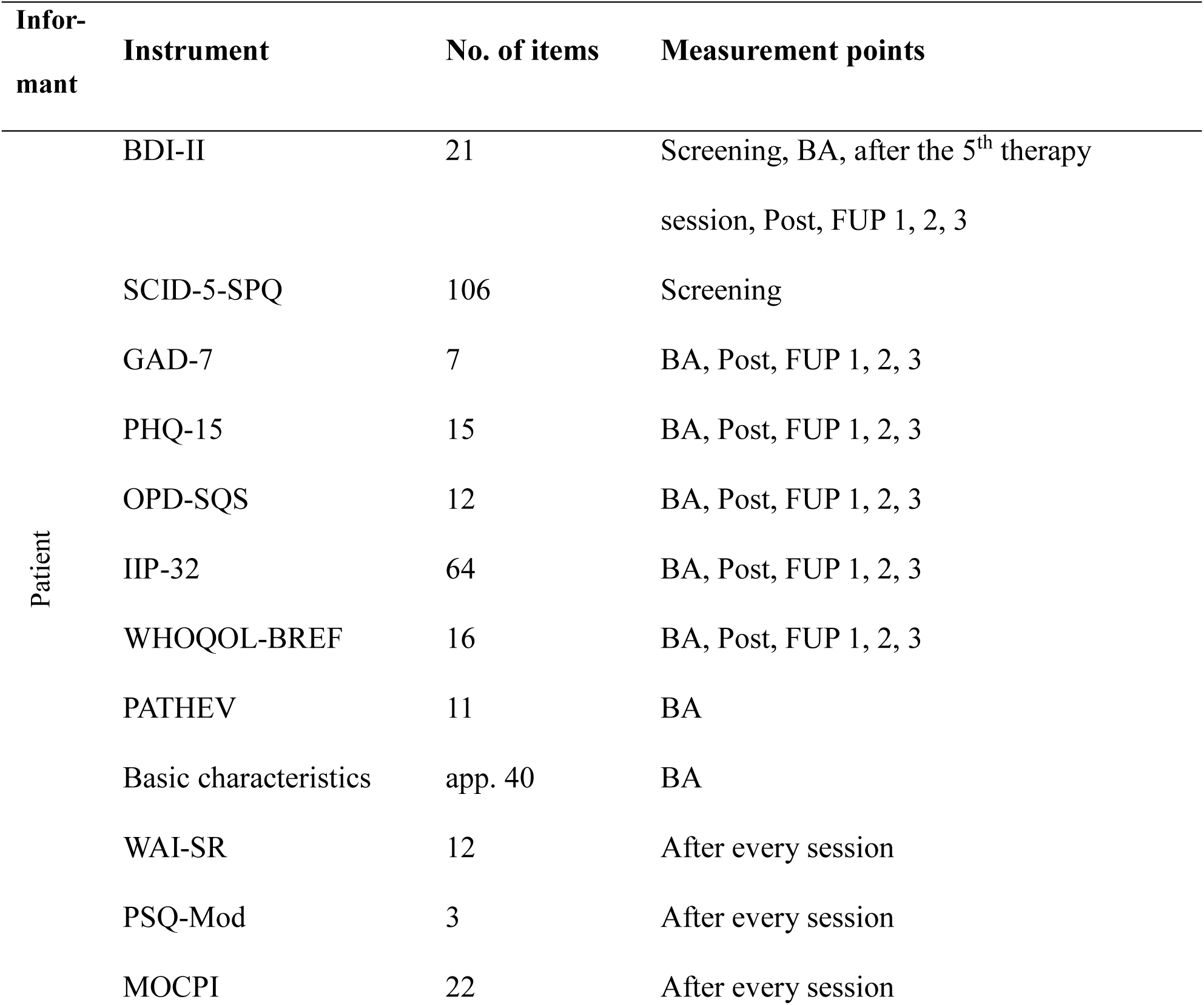
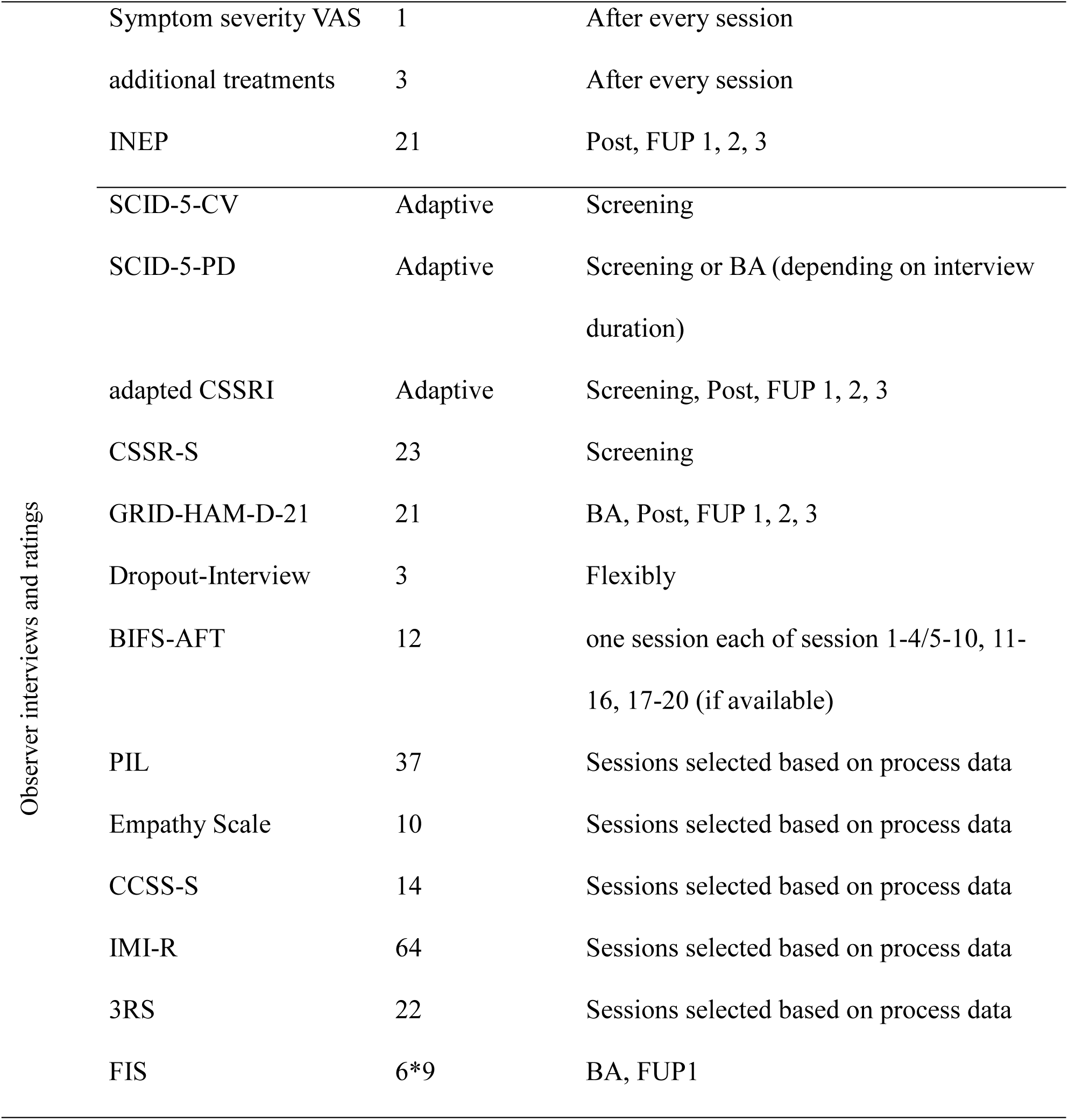
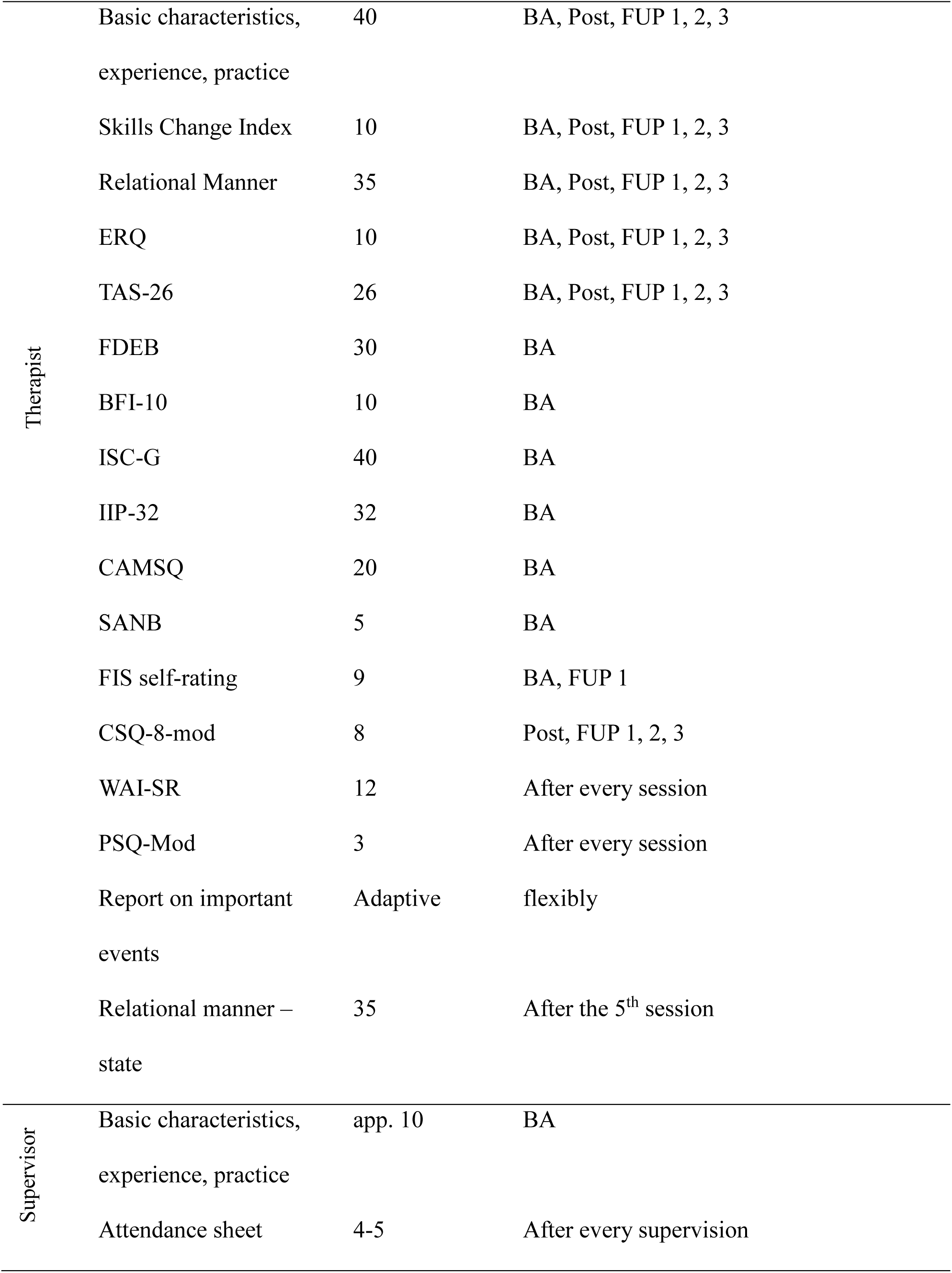
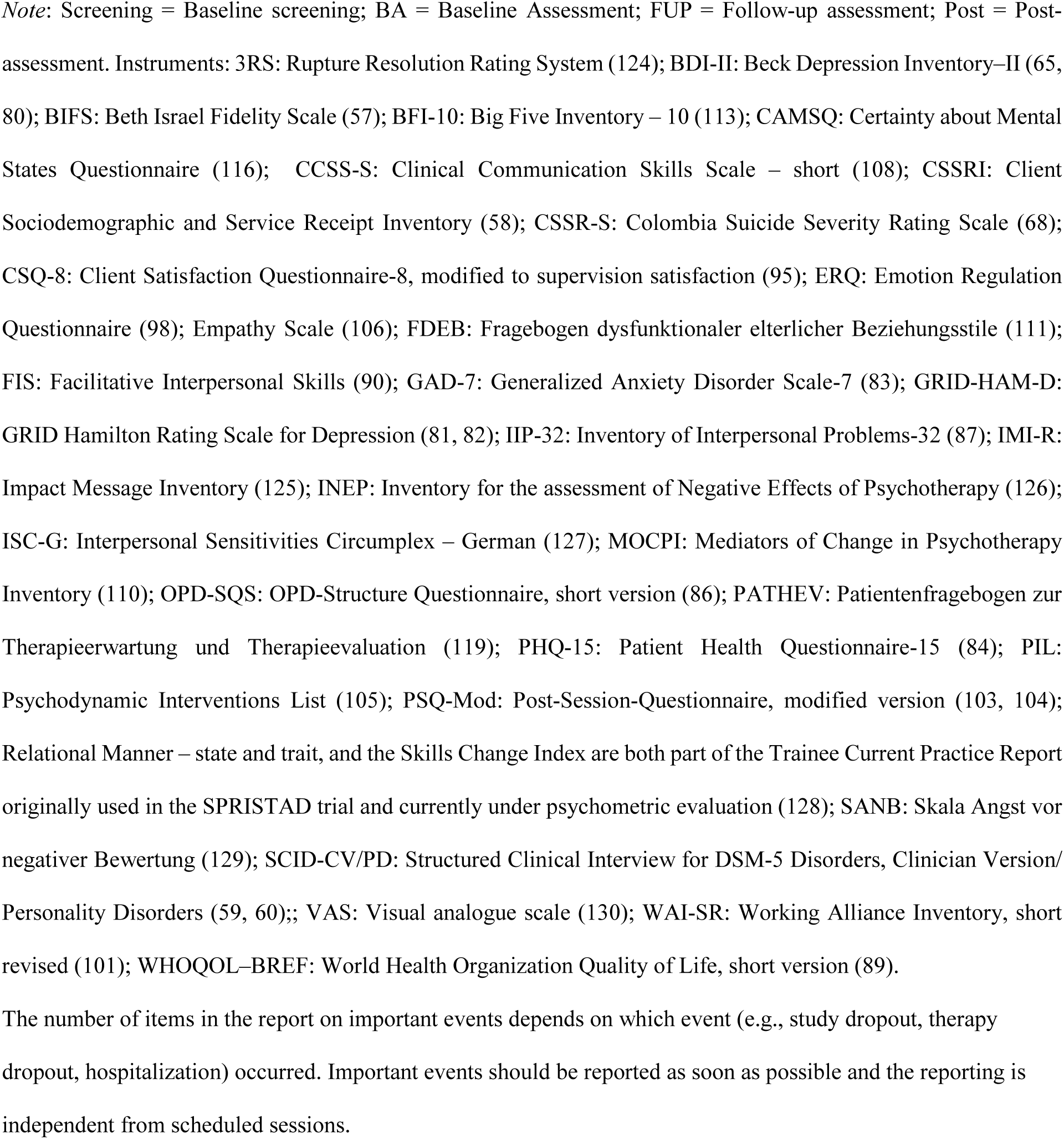
Data and measurement time points.

### Statistical analyses

Results are reported according to the CONSORT statement (131). Analyses are carried out with standard software: SPSS, Version 29 or newer (Armonk, NY: IBM Corp.), SAS, Version 9.4 or newer (Cary, NC: SAS Institute Inc.), or R (Version 4.3.3). A statistical analysis plan that contains the details of all planned analyses will be finalized before database lock.

Descriptive data are presented overall and separated by randomized group. As descriptive statistics, mean, standard deviation, median, first and third quartile or minimum and maximum are calculated for continuous variables. For categorical variables, absolute and relative frequencies are reported.

First primary outcome (hypothesis I): We use a linear mixed model with repeated measures with changes from baseline as dependent variable, intervention group, therapeutic approach (CBT vs. PDT), time and comorbid SCID-5 personality disorder (yes vs. no) as fixed effects, baseline BDI-II score as covariate, time by intervention group interaction, and cooperation center, supervision group (if possible), therapist (= cluster) and patient as nested random effects. The interaction term is eliminated from the model if the corresponding interaction p-value is >0.15. The contrast of the intervention group after 20 weeks is the result of the primary analysis. This result is considered in a confirmatory manner according to the approach described below.

Second primary outcome (hypothesis II): We perform a mixed logistic regression with the binary variable patient dropout after 20 weeks (yes vs. no) as dependent variable, intervention group, therapeutic approach (CBT vs. PDT), and comorbid SCID-5 personality disorder (yes vs. no) as fixed effects, and cooperation center, supervision group (if possible), and therapist as nested random effects.

The two primary hypotheses are hierarchically ordered and tested one after the other at the two-sided 5% level. Hypothesis II is only tested if hypothesis I could be rejected. If hypothesis I cannot be rejected, hypothesis II is no longer examined confirmatory but exploratory. This procedure ensures a family wise type I error of 5%.

These analyses will be conducted in the full analysis set which is as complete as possible and as close as possible to the intention-to-treat ideal of including all randomized participants [120]. Missing values are treated by direct maximum-likelihood imputation to allow an intention to treat analysis, which results in unbiased estimations under the missing-at-random assumption. Missing values for treatment dropout can only be due to both patient and therapist leaving the study.

Sensitivity analysis for primary endpoints: To investigate the robustness of the results of the primary analyses we will conduct sensitivity analyses with multiple imputation by chained equation as imputation strategy. Additionally, primary analyses are repeated in the per protocol (PP) sample. Definition PP sample: patients who receive therapy by trained or non-trained and accordingly supervised therapists as intended by their group allocation without major protocol deviations (i. e., at least 12 sessions within 20 weeks for BDI-II outcome analysis; no changes with respect to psychopharmacotherapy, additional treatments as allowed for BDI-II outcome and treatment dropout) and who complete the assessments within the defined time frames.

Secondary outcomes: Secondary outcomes are examined exploratory with analogous methods appropriate for the scale level (i.e. mixed linear, logistic, ordinal, or poisson/negative binomial regression). The contrasts of the intervention vs. control group at week 5, 10, 15, 35 of BDI-II changes from baseline of the model of the first primary outcome are used as results of secondary outcome analysis.

Mediators: In path models, we test the hypotheses that patient’s outcome is mediated by the average level of therapeutic alliance across sessions (WAI patient-rated); ratio of unresolved/ resolved alliance ruptures (PSQ-Mod patient-rated); convergence of patient’s and therapist’s alliance ratings over time (WAI patient-/therapist-rated); therapists’ usage of interventions referring to the therapeutic relationship in the here and now; therapists’ usage of nondirective supportive techniques (validation, expressing emotional sympathy, repeating paraphrizing, summarizing; PIL); therapists’ competence in session (empathy); therapists’ adherence to AFT techniques (BIFS-Subscale AFT).

Additional analyses: a) To test whether MAFT-D is equally effective in CBT and PDT, we examine the interaction between intervention group and therapeutic approach. Prior to this analysis, we examine baseline differences (depression severity, chronicity, comorbidity) between therapeutic approaches (CBT/PDT). Variables for which we detect clinically relevant group differences will be added as covariates. Cooperation center will be included as random effect. b) We examine the adherence measure BIFS (adherence with AFT-techniques, observer-rated) as an outcome, with the same approach as in primary analyses (extent of contamination bias). c) Safety endpoints are determined using frequency tables and if possible (mixed) logistic or linear regressions to compare the event frequencies/groups. d) Subsidiary research question concern predictors, moderatos, and/ or mediators (patient-, therapist-, patient-therapist match- and process-related variables) of a) therapeutic macro and micro outcomes (e.g., dropout, alliance), b) the effectiveness of MAFT-D (e.g., presence of a personality or therapeutic orientation as moderators of MAFT-D effectiveness), c) observer-rated interpersonal therapeutic skills and skills development and d) the successful resolution of alliance ruptures.

Interim analyses are not planned. For secondary outcomes and analyses, missing values are not imputed and no adjustment for multiple testing is conducted.

### Ethics and dissemination

The study is being conducted in compliance with the protocol, Good Clinical Practice, the declaration of Helsinki and the CONSORT statement (131). All study patients, therapists and supervisors need to provide written informed consent to the main study team prior to inclusion. Study participation is completely voluntary and can be cancelled at any time without negative consequences. Patients who terminate study participation can continue their treatment. The study was approved by the Ethics Committee of the Psychologische Hochschule Berlin (EK2024/11). Additionally, we obtained secondary votes in each federal state where one or more study centers are located. Any major modifications to the study procol will be covered by amendments and communicated to the relevant parties (e.g., Data Safety and Monitoring Board, DSMB, see below). All personal data collected as part of the study, following the consent of the study participant, are subject to confidentiality and the provisions of the General Data Protection Regulation. All questionnaire, interview, and rating data are collected pseudonymously within the secuTrial eCRF and REDCap surveys. This means that the participant’s name and all direct identifiers are replaced with a pseudonym making re-identification outside of the study no longer possible. Access to the code key list that enables the association between study data and personal information of the study participant is limited to study staff explicitly authorized by the principal investigator. The code key list is stored securely and separately from all other study data (e.g., questionnaire data). The individual code key will be permanently erased following the completion of data collection and after the obligatory archiving time frame.

There is subject insurance for study patients. They receive the insurance confirmation and insurance conditions together with the study information. Assessments might be experienced as time-consuming. We do not expect other risks or adverse events due to study participation in patients or therapists. Positive effects resulting from treatments in both groups can be expected. There is no placebo or waiting list group. Both evidence-based treatments will be conducted according to state-of-the-art CBT/PDT and will be supervised by certified experienced psychotherapists. Frequency and dose of supervision corresponds to usual standards.

Results will be published in peer-reviewed scientific journals following the ICMJE guidelines and disseminated to the general public, patient organisations and media.

Public Involvement: Implications and recommendations for psychotherapy studies and further psychotherapy training are to be derived from the project results. These proposals will be discussed with relevant stakeholders (e.g., the clinical directors of the training institutes).

### Quality Assurance and Safety

During the trial, quality control and quality assurance will be ensured through central monitoring based on the risk assessment of the study according to ICH Good Clinical Practice (GCP). The Clinical Trial Office (CTO) will perform coordination, implementation, and conduction of monitoring at the main study center following its applicable Standard Operating Procedures (SOPs). Monitoring will be performed according to SOPs and study specific requirements in accordance with the patient recruitment. The monitoring strategy will be defined in a monitoring plan with special attention to critical data and processes. The verification will focus on the key study data, e.g. signed informed consent, adherence to inclusion and exclusion criteria and participant safety or adverse events. All the other data are checked on the basis of a representative sample, e.g. documentation on primary objectives. Unclear and incomplete data will trigger increasing in-depth monitoring of the respective data.

Data management creates a data validation plan, containing additional logic checks for plausibility and consistency not available within the electronic case report forms (eCRF). Query analysis will be performed in SAS on an up-to-date export from the CRF. Found inconsistencies, implausible or missing data will be flagged by a data management query within the CRF. A possibly false information can be corrected through directly changing values within the eCRF or must be flagged as accepted failures or false positives. Changes within the documented information will be tracked via the audit trail, which is a core function of the eCRF.

After the last participant had its last visit, all queries and all centers were closed, the database will be locked. Electronic documentation includes all exported files (SPSS, SAS, CSV, EXCEL), SAS scripts, data protocol and the closed database.

An independent DSMB will be regularly provided with all safety aspects of the trial and will review the safety data. Members of the DSMB are: Prof. Dr. Christiane Steinert, International Psychoanalytic University, Berlin, licensed psychotherapist; Prof. Dr. Elmar Brähler, Department of Psychosomatic Medicine and Psychotherapy, University Medical Center Mainz, experienced researcher and DSMB member in former psychotherapy trials, and Dr. Theresa Keller, Charité - Universitätsmedizin Berlin, independent statistician. At annual intervals, a meeting of the board will be scheduled to review whether the recruitment plan is on target, to ensure compliance with ethical principles, adherence to protocol, to check data quality and accuracy, and to advise whether to continue, modify, or stop the trial. The DSMB will serve as link to the funding organisation and provide them with information and advice. In the event of a respective advice from the DSMB, the steering committee will finally decide whether to stop the trial (members: Prof. A. Gumz, PI; Prof. A. Zapf, statistician, Prof. S. Singer - Institute for Medical Biostatistics, Epidemiology and Informatics, University Medical Center of the Johannes Gutenberg University Mainz, independent member, and Prof. L. White - Clinical Child and Adolescent Psychology and Psychotherapy, Bremen University, independent member).

Important events (e.g., therapy dropouts, hospitalization) can be reported by the therapist at any time. The study team will be alerted about incoming reports via email. Supervisors will be available at all study centers to decide about crisis interventions (e.g., need for referrals). We will assess side effects/ risks with questionnaires and interviews.

## Discussion

This study protocol presents a comprehensive and innovative project designed to examine the impact of MAFT-D compared to psychotherapy training and treatment-as-usual (TAU) for trainee therapists and their patients with depressive disorders.

### Strengths of the study

The proposed project focuses on one of the most prevalent disorders, is dedicated to a topic highly relevant to everyday practice, and uses a randomized, controlled multi-center design including about 16 training institutes across Germany. Thus, the results have great potential to substantially contribute to an evidence-based psychotherapy training and improved outcomes for patients with depression. By involving numerous institutes, the study not only ensures generalizability within a naturalistic setting, but it could also directly impact clinical training, especially if positive results lead to the integration of MAFT-D into psychotherapeutic training curricula. The intervention itself has a transtheoretical nature and could be incorporated into various existing training and continued education programs at comparatively low cost (132).

Notably, AFT has so far been more widely studied in the context of personality disorders (42, 133), with limited evidence available for patients with depression. This study focuses on patients with depression, but includes those with comorbid personality disorders, stratifying for this variable. Thus, the study also stands to clarify potential differential effects of MAFT-D depending on patients’ psychopathological characteristics, contributing to a more nuanced understanding of alliance-focused approaches across various disorders.

Moreover, the study provides valuable insights into the active ingredients of psychotherapy for depression. The chosen design allows for the consideration and differentiation of patient and therapist effects, as well as various common factors measured from different perspectives (patients, therapist, observer). The knowledge generated may contribute to a better understanding of change mechanisms in psychotherapy overall, which constitutes a critical step toward more effective treatments.

### Methodological considerations and potential challenges

We aim for a rigorous methodology and different study design components are implemented to effectively minimize bias: Randomization, blinding of raters, prevention of contamination, control of therapy allegiance and of overlapping treatments as well as for confounding factors are described and warranted by corresponding measures. Additionally, the sample size is sufficiently large to enable generalizable findings across broader populations with depressive disorders.

The study’s hierarchical testing framework first assesses symptom reduction, followed by dropout rates. While this sequence reasonably suggests that dropout rate effects might be less relevant without significant symptom reduction, we would like to point out that one could also argue that both hypotheses hold independent value and could merit separate evaluation, given their relevance to treatment quality.

The central aim of this study is to examine how therapist training affects therapy outcomes for patients. This means that the primary outcomes are measured not in those who receive the intervention but in the patients they treat. Although this approach might seem self-evident in the context of therapist trainings, few studies actually adopt it. Most evaluations of therapist trainings focus instead on therapist outcomes, such as improvements in therapeutic skills (e.g. (43)). While one could argue that improved therapeutic skills should lead to better therapy outcomes, this remains an assumption rather than a proven fact. Therefore, this study was designed to assess both changes in therapeutic skills (as perceived by observers and therapists themselves) and the more essential patient outcomes.

### Limitations

A key methodological aspect to consider is the supervision frequency, set at one session for every four therapy sessions. While this represents the common standard and legal requirement for trainee therapists in our country, this frequency might deviate from other clinical trials or the clinical practice in others regions. The supervision frequency may influence outcomes but also reflects local real-world practice conditions, adding ecological validity to the study.

For the purpose of comparability with other studies, the primary outcome was set to be measured at 20 weeks. However, this introduces a risk that the intervention dose may be insufficient by this time point, and effects might only become apparent at later follow-ups.

Another limiting factor is that, due to the large number of cooperating psychotherapy training institutes and the relatively small number of participating therapists per institute (typically eight), it remains unclear whether we will be able to disentangle the effects of the center from those of the supervisor or supervision group. If feasible, we plan to include supervision group as a random effect.

Additionally, it might have been valuable to record not only therapy sessions but also supervision sessions. This could have provided deeper insights into adherence to MAFT-D and rupture-repair processes occurring during supervision. Unfortunately, this was not feasible within the scope of the current study.

Finally, selective attrition poses a risk: if MAFT-D indeed reduces dropout rates, this might lead to differences in attrition between the intervention and control groups. This is particularly relevant to the analysis in the intention-to-treat population using the mixed linear model, which leads under the “Missing-at-Random” (MAR) assumption—that missing data occur at random rather than systematically (134). Various mechanisms are implemented to prevent study dropout in patients who drop out of therapy (e.g., incentives). However, this aspect will require careful consideration and control.

### Conclusion

In summary, this protocol presents an innovative and carefully designed study to test the effectiveness of MAFT-D for trainee therapists and their patients with depression. The study holds the potential to significantly enhance the quality of psychotherapy training and to contribute to a deeper understanding of mechanisms of change in psychotherapy. With positive findings, the study could have immediate implications for psychotherapeutic education and clinical practice. Despite some limitations and methodological challenges, the study’s naturalistic approach underscores its relevance and ecological validity, positioning it to make a meaningful contribution to improving treatment outcomes and reducing dropout rates in depression therapy.

## Acknowledgments

We would like to thank all current and former members of our research group who contributed to our research described in this article. In particular, we would like to thank Jelka Berger, Merle Longley, Dr. Christopher Marx, Dr. Thomas Munder, Dr. Kai Rugenstein, Carina Schlipfenbacher, Kyra Toussaint, Lena Walther, and Miriam Warmuth. We also thank our partners at the cooperating training institutes with whom we have worked together or who have agreed to cooperate in the future.

## Statement of Ethics

The study was reviewed and approved by the Ethics Committee of the Psychologische Hochschule Berlin (AZ: EK2024/11). Secondary votes were obtained for each federal state where a study center is located. Written informed consent was obtained from all study participants (patients, therapists, supervisors).

## Data availability

On request, anonymized raw data will be made available to scientific colleagues after finishing the trial. During the publication process, relevant trial data will be uploaded to the Dryad Digital Repository.

## Author Contributions

AG formulated the research question and conception of the study. AG, DK, LR, AZ and AD designed the study. CE and CM developed the AFT, provided their original materials and supported the research team with information and their experience. TA contributed to the implementation of measurement of therapeutic competence (Facilitative interpersonal skills) as an outcome und predictor variable by providing his original materials and supporting the research team with information and his experience. RS, GS, LS and LRo supported the study team with their experience in data collection and management and in the implementation of the clinical study. AG tested the described MAFT-D procedures in clinical context. UW provided a questionnaire. AG, DK, LR, CMM, KE, UW, and FJ contributed to the recruitment of cooperating training institutes. AG drafted the manuscript. DK, AZ, and AD provided input to the first draft. All authors revised it critically for important intellectual content. All authors gave their final approval of the version to be published. All authors agreed to be accountable for all aspects of the work in ensuring that questions related to the accuracy or integrity of any part of the work are appropriately investigated and resolved.

